# The Effects of Maternal Obesity on Some Obstetric Features: A Comparative and Descriptive Study

**DOI:** 10.1101/2023.05.24.23290497

**Authors:** Merve Ekiz, Aysegul Durmaz

**Affiliations:** An Emergency Department, Izmir Provincial Directorate of Health Gaziemir Nevvar Salih Isgoren State Hospital, Izmir, Türkiye; Department of Midwifery, Faculty of Health Sciences, Kutahya Health Sciences University, Kutahya, Türkiye

**Keywords:** maternal, obesity, weight, body mass index, postpartum

## Abstract

**Background:** The worldwide increase in obesity triggers the increase in the incidence of obesity during pregnancy. In this study, it was aimed to examine the effects of maternal obesity on some obstetric features.

**Methods:** This study was conducted in a tertiary hospital with a comparative group, crosssectional and descriptive design. The data of the study were collected with a Data Collection Form created by the researchers. Descriptive statistics, Pearson’s chi-squared test, independent-samples t-test, and multinomial regression analysis were used to analyze the data.

**Results:** In this study, women were divided into two groups, namely the obese group (BMI>29.9 kg/m^2^) and the normal weight group (BMI 18.5-24.9 kg/m^2^). There was no significant difference between the groups in terms of their height, education level, employment status, income level, and place of residence (p>0.05). It was determined that the obese group had higher rates of multiparity, postmature deliveries, and fetal distress developing in their babies on labor (p<0.05). There was no significant difference between the groups in terms of their modes of delivery, onset of labor, and the requirement of interventions during the second stage of the labor (p>0.05). In the logistic regression analysis, maternal obesity was found to increase the rates of multiparity by 1.758 times (1.038-2.978 CI, p=0.036), post-maturity by 5.902 times (1.283-27.164 CI, p=0.023), and moderate postpartum hemorrhage by 2.286 times (1.433-3.646 CI, p=0.001).

**Conclusion:** It is important that women who have obesity problems in the preconception period are counseled to help them reach a normal BMI. Health care professionals should advise women on healthy nutrition and weight control during both the pregnancy and postpartum periods.

## Introduction

Obesity is a common, serious, and costly disease [1]. The term obesity is used for greater weight gain than is generally considered healthy for a given height [2,3]. A body mass index (BMI) of 30 kg/m^2^ or above in pregnancy at the time of first prenatal follow-up is defined as maternal obesity [4]. BMI is used to determine overweight or obesity [5]. BMI is a person’s weight (in kg) divided by their height in meters squared (BMI=Kg/m^2^). WHO defines women who have a BMI of 25 kg/m^2^ or above as overweight, and women with a BMI of 30 kg/ m2 or above are defined as obese.

Globally, there are more obese people than underweight people. This occurs in all regions except sub-Saharan Africa and some parts of Asia. In WHO regions, the rate of BMI≥30 kg/m2 was determined as 29% in the Americas, 25.3% in Europe, 19.5% in the Middle East, and 4.6% in South-East Asia. The proportion of women with BMI values equal to or greater than 30 kg/m^2^ was reported to be 31.7% in the USA, 27.1% in Europe, 24.3% in the Middle East, and 6% in South-East Asia [6]. In the 2018 report of the Turkey Demographic and Health Survey, it was specified that 29% of women aged 15-49 were overweight, and 30% were obese [7]. In parallel with the increase in obesity in the general population, the incidence of obesity in pregnancy increases worldwide [8]. A study which examined the most recent version of the four national population guidelines reported an increase in the rates of obesity in pregnant women in parallel with the increase in the general population [9]. The Royal College of Obstetricians and Gynaecologists reported that 21.3% of the UK population is obese, and less than half (47.3%) of pregnant women have a normal BMI [10]. In a study conducted in Australia, it was found that 55% of pregnant women were overweight or obese, and 43.6% gained more weight than recommended during pregnancy [11].

Obesity in pregnancy is considered a high-risk condition as it is associated with many complications [12]. Health risks for obese pregnant women were reported in a systematic review. These included gestational diabetes, gestational hypertension, preeclampsia, antenatal anxiety, and mental health problems including depression and postpartum depression, increased rates of cesarean section, instrumental delivery, and increased rates of post-cesarean section surgical site infections [13]. It is known that obese pregnant women who gain less than 5 kg of weight during pregnancy have a decrease in the incidence of risky conditions such as hypertension, eclampsia, gestational diabetes, and cesarean section. It was observed that there was an increase in maternal risks in obese pregnant women who gained weight by over 9 kg during pregnancy [14]. The relevant literature shows that maternal obesity negatively affects maternal health during the pregnancy, childbirth, and postpartum periods. In this context, it was aimed in this study to determine the effects of maternal obesity on some obstetric features. Based on the literature, the study sought answers to two research questions:

1. What are the effects of maternal obesity on the obstetric characteristics of women?
2. What are the relationships between maternal obesity and the obstetric characteristics of women?

## Materials and methods

### Study design and setting

This study was conducted in a tertiary hospital with a comparative group, cross-sectional and descriptive design. The data of the study were obtained between 15 June and 31 December 2021. The research was carried out in one of the most populated districts of Turkey’s 3rd largest city. There are approximately 5400 births per year in this district.

The population of the study consisted of women who gave birth in the hospital where the study was conducted, who had a BMI in the range of 18.5-24.9 kg/m^2^ before pregnancy and those in the early postpartum period with a BMI higher than 29.9 kg/m^2^. The minimum sample size of the study was calculated by power analysis. According to the power analysis, a margin of error (α) of 0.05, 95% power to represent the population, and with an effect size of 0.448, the required sample size was determined as 262 women, including 131 women with obesity problems and 131 women with normal weight [15]. When the effect size was taken as 0.383, the error margin (α) was 0.05, and power to represent the population (confidence interval) was 95%, the required sample size was calculated as 298 women, including 149 obese women and 149 normal-weight women [16]. A total of 320 women, 160 women in each group (case and control groups), were included in the study by considering the possibility of loss of data during the conduct of the study. The study was completed with 310 women. 155 women with a BMI of >29.9 kg/m^2^ and 155 with a BMI in the range of 18.5-24.9 kg/m^2^ were included in the study. In the selection of the sample, the purposive sampling method was used. In the post hoc power analysis of the study, the 1-β value was calculated as 0.99.

Purposeful sampling technique was used in this study to reach obese women in the puerperal period. Not using the randomization method increased the risk of bias. However, the study was descriptive and included a specific group (maternal obese women).

### Participants

Pregnant women who volunteered to participate in the study, lived in Burdur province, used herbal products in their current pregnancy, were not diagnosed with a psychological disease and not receiving any medications, were older than 18 years, were literate in Turkish, and completely responded to the data collection forms were included in the study. Pregnant women who did not use herbal products were excluded from the study. Also, pregnant women who did not meet the inclusion criteria were excluded from the study.

Those who gave birth in the hospital, were over the age of 18, could read, write, and speak Turkish, and didn’t have any chronic diseases (e.g., diabetes, hypertension, thyroid diseases), had pre-pregnancy weight values that could be accessed from their medical records (at the family health center or hospital), agree to participate in the study, a pre-pregnancy BMI value of 18.5-24.9 kg/m^2^ or >29.9 kg/m^2^ depending on their allocation group, were in the first 24 hours of the postpartum period, and did not have physical or mental health problems that would prevent their participation in the study were included in the study. Women who had a BMI range of 26.0-29.9 kg/m^2^ before pregnancy and those who wanted to leave the study at any stage of the study were excluded.

### Data Collection Tool

The data were obtained with the “Data Collection Form” developed by the researchers in line with the relevant literature.

*Data Collection Form* consisted of questions about the sociodemographic characteristics (age, current body weight, pre-pregnancy body weight, total weight gained during pregnancy, height, pre-pregnancy BMI, postpartum BMI, postpartum waist circumference, education, family type, employment status, income level, place of residence) and obstetric characteristics (parity, time of childbirth, mode of delivery, labor induction status, status of having an intervention in the second stage of birth, status of fetal distress development in childbirth, status of hemorrhage in the emergency puerperium period) of the participants [13,17-18].

### Data collection procedure

According to the classification used in the studies conducted by WHO (2021), CDC (2019), and NICE (2014), women with a BMI of 30 kg/m^2^ or greater are obese, and those with a BMI in the range of 18.5-24.9 kg/m^2^ accepted to have a normal weight [2,5-6]. The women included in the study were divided into two groups as obese group and normal weight group. The purpose of the study was explained to the women included in the study and the consent form was signed. The researcher obtained the data by face-to-face interview technique. The physical examinations and measurements of the women who were mobilized within the first 24 hours postpartum were performed, and the necessary information was recorded in the forms. Additionally, the information given by the participants was confirmed by examining their hospital record files. Missing data were completed based on information obtained from these files and from the family health centers where the participants were registered. Filling out the data collection forms, making the measurements, and conducting the physical examinations took approximately 20-25 minutes for each participant.

Opinions and suggestions of five midwife with Ph.D were taken about the data collection form developed by the researchers to ensure its validity and applicability. After the final of the data collection form was given, a pretest was applied to 10 pregnant women (5 experimental and 5 control groups). Pre-tested pregnant women were excluded from the study. While collecting the data of the study, participants were excluded from the study because 3 women refused to participate in the study (MO:2, NW:1), 3 participants were under the age of 18 (MO:1, NW:2), 2 participants were illiterate (MO:1, NW:1), 1 participant could not speak Turkish (NW:1), 1 participant did not want to continue the study (MO:1) (Figure 1).

**Fig 1.**
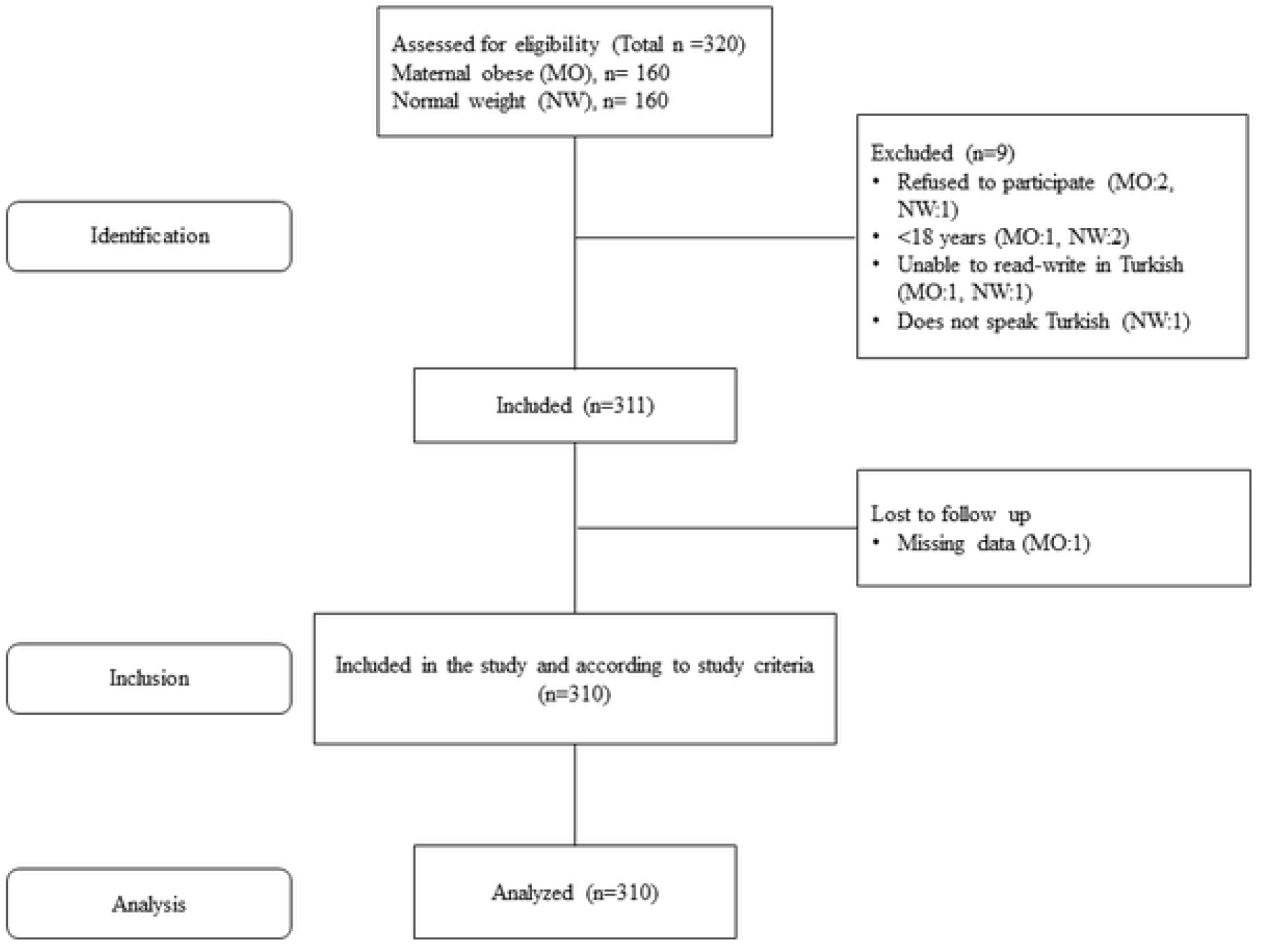
Follow diagram of study.

### Data analysis

The analysis of the data obtained in the study was conducted with appropriate statistical methods in the IBM SPSS Statistics 22 (SPSS Inc., Chicago, IL) program. Descriptive statistics (mean, standard deviation, frequency, percentage, and distribution range) were used to analyze the main information. The normality of the distribution of the data was tested by skewness values, kurtosis values (-1.5-+1.5), and the Kolmogorov-Smirnov test. Parametric tests were used to analyze the data, as the data were normally distributed. The independent-samples t-test, Pearson’s chi-squared test, and multinomial regression analysis methods were used in the analyses. *p*<.05 was considered significant in the interpretations of the results of the analyses.

### Ethical aspect of the study

The approval of an Ethics Committee (Decision No: 2021/08-19), the permission of the institution, and permissions for use of the scales were obtained to conduct the study. The purpose of the study was verbally explained to the potential participants, and an informed consent form was signed by those who agreed to participate in the study. The principles of the Declaration of Helsinki (revised in Brazil 2013) were followed in the research protocol. The study data were stored within the scope of personal data protection principles. Due to anonymity and confidentiality, no personally identifiable data such as name and address were obtained. No women were harmed as a result of this study.

## Results

In this study, the participants were divided into two groups: maternally obese (MO:BMI >29.9 kg/m^2^) and normal weight (NW:BMI 18.5-25.9 kg/m^2^) groups. The study was completed with a total of 310 puerperium participants, 155 of whom were maternal obese and 155 with normal weight (Figure 1). Therefore, their pre-pregnancy weight (MO: 83.50±9.87 kg vs NW: 56.38±7.15 kg), current weight (MO: 91.14±11.30 kg vs NW: 67.33±8.97 kg), total weight gained during pregnancy (MO: 11.08±4.75 kg vs NW: 12.33±6.03 kg), BMI pre-pregnancy (MO: 32.53±2.77 kg/m^2^ vs NW: 21.85±2.06 kg/m^2^), postpartum BMI (MO: 35.72±3.57 kg/m^2^ vs NW: 25.96±3.14 kg/m^2^), and postpartum waist circumference (MO: 107.06±9.28 cm vs NW: 90.97±8.66 cm) variables were significantly different (p<0.05). Additionally, there was a significant difference between the groups in terms of their mean ages (MO: 28.78±6.14 vs NW: 26.89±6.00) and family types (nuclear family; MO: 69.0% vs NW: 82.6%) (p<0.05). The mean heights of the participants (MO: 159.95±4.42 cm vs NW: 160.92±5.08 cm), their education levels (primary education; ML: 43.9% vs NW: 45.2%), and employment statuses (not working; ML: 93.5% vs NW: 92.3%) were not significantly different (p>0.05). There was also no significant difference between the groups in terms of their income levels (income less than expenditures; MO: 69.0% vs NW: 69.7%) or places of residence (province; MO: 71.0% vs NW: 78.7%) (p>0.05) (Table 1).

**Table 1.**
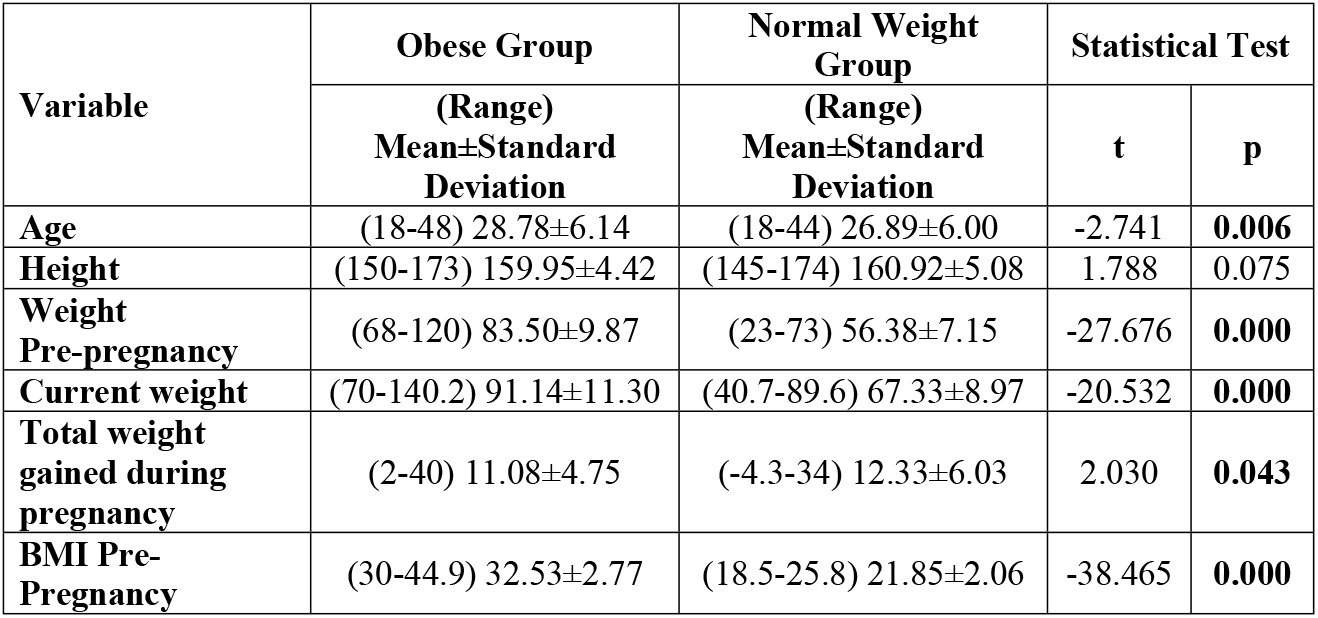
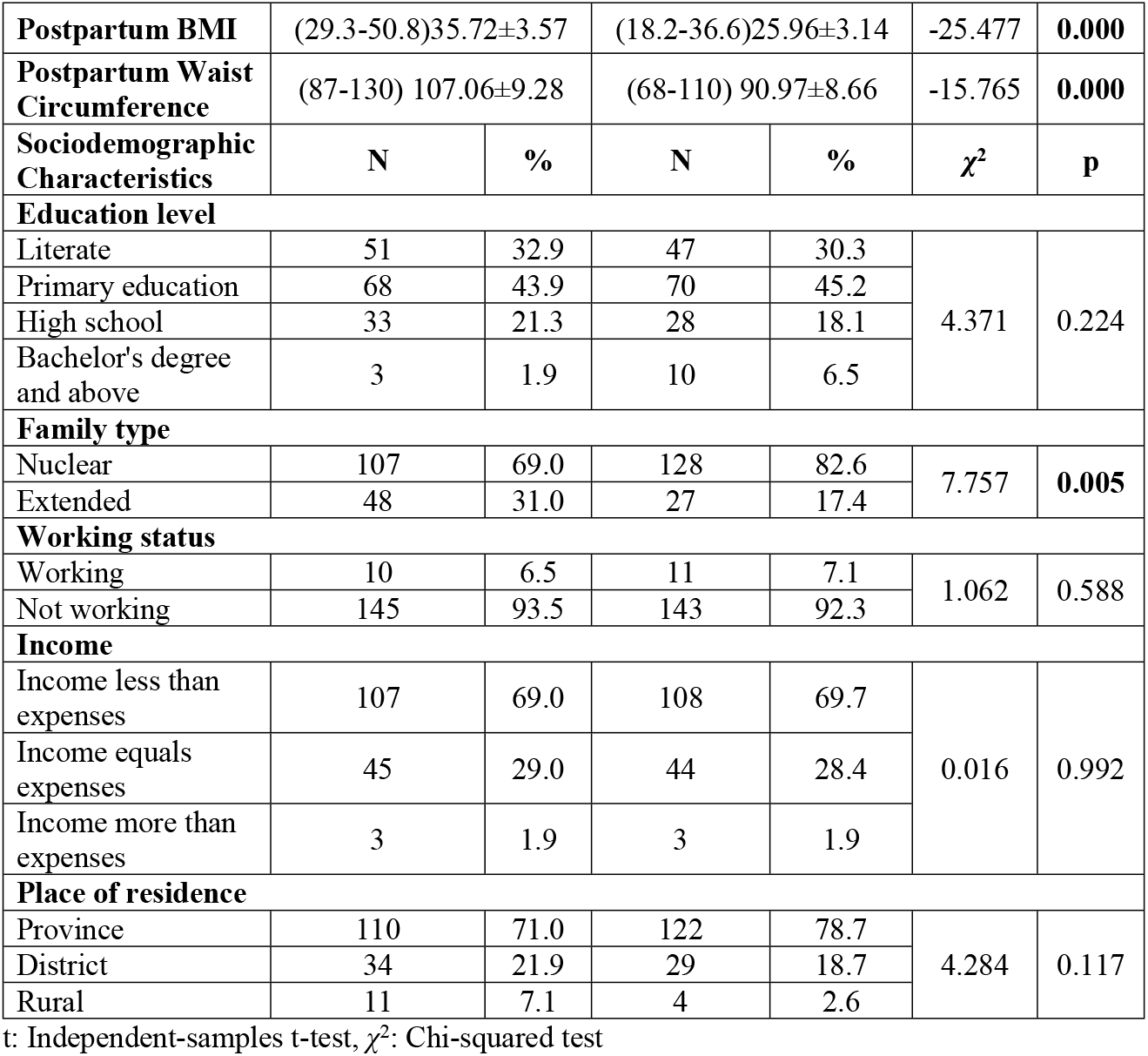
Socio-demographic characteristics of the women in the groups (N=310)

| Variable | Obese Group | Normal Weight Group | Statistical Test |  |
| --- | --- | --- | --- | --- |
|  | (Range)<br>Mean±Standard Deviation | (Range)<br>Mean±Standard Deviation | t | p |
| Age | (18-48) 28.78±6.14 | (18-44) 26.89±6.00 | -2.741 | **0.006** |
| Height | (150-173) 159.95±4.42 | (145-174) 160.92±5.08 | 1.788 | 0.075 |
| Weight Pre-pregnancy | (68-120) 83.50±9.87 | (23-73) 56.38±7.15 | -27.676 | **0.000** |
| Current weight | (70-140.2) 91.14±11.30 | (40.7-89.6) 67.33±8.97 | -20.532 | **0.000** |
| Total weight gained during pregnancy | (2-40) 11.08±4.75 | (-4.3-34) 12.33±6.03 | 2.030 | **0.043** |
| BMI Pre-Pregnancy | (30-44.9) 32.53±2.77 | (18.5-25.8) 21.85±2.06 | -38.465 | **0.000** |

|  |  |  |  |  |  |  |
| --- | --- | --- | --- | --- | --- | --- |
| Postpartum BMI | (29.3-50.8)35.72±3.57 |  | (18.2-36.6)25.96±3.14 |  | -25.477 | 0.000 |
| Postpartum Waist Circumference | (87-130) 107.06±9.28 |  | (68-110) 90.97±8.66 |  | -15.765 | 0.000 |
| Sociodemographic Characteristics | N | % | N | % | χ<sup>2</sup> | p |
| Education level |  |  |  |  |  |  |
| Literate | 51 | 32.9 | 47 | 30.3 | 4.371 | 0.224 |
| Primary education | 68 | 43.9 | 70 | 45.2 |  |  |
| High school | 33 | 21.3 | 28 | 18.1 |  |  |
| Bachelor's degree and above | 3 | 1.9 | 10 | 6.5 |  |  |
| Family type |  |  |  |  |  |  |
| Nuclear | 107 | 69.0 | 128 | 82.6 | 7.757 | 0.005 |
| Extended | 48 | 31.0 | 27 | 17.4 |  |  |
| Working status |  |  |  |  |  |  |
| Working | 10 | 6.5 | 11 | 7.1 | 1.062 | 0.588 |
| Not working | 145 | 93.5 | 143 | 92.3 |  |  |
| Income |  |  |  |  |  |  |
| Income less than expenses | 107 | 69.0 | 108 | 69.7 | 0.016 | 0.992 |
| Income equals expenses | 45 | 29.0 | 44 | 28.4 |  |  |
| Income more than expenses | 3 | 1.9 | 3 | 1.9 |  |  |
| Place of residence |  |  |  |  |  |  |
| Province | 110 | 71.0 | 122 | 78.7 | 4.284 | 0.117 |
| District | 34 | 21.9 | 29 | 18.7 |  |  |
| Rural | 11 | 7.1 | 4 | 2.6 |  |  |

t: Independent-samples t-test, $\chi^2$ : Chi-squared test

The majority of the participants in both groups were multiparous (MO: 80.6% vs NW: 70.3%) and gave birth at term (MO: 79.4% vs NW: 85.2%). Additionally, post-mature births were seen significantly more commonly in the obese group (MO: 7.1% vs NW: 1.3%) (p<0.05). There was no significant difference between the groups in terms of their modes of delivery, their statuses of receiving labor induction, and their statuses of undergoing any intervention status in the second stage of labor (p>0.05). It was observed that fetal distress at birth developed only in the infants of the participants in the obese group (MO: 20.6% vs NW: 0.0%), and moderate hemorrhage was observed significantly more frequently in the obese group (MO: 51.6% vs NW: 32.3%) (p<0.05) (Table 2).

**Table 2.** Obstetric characteristics of the women in the groups (N=310)

| Variable | Obese Group | | Normal Weight Group | | $\chi^2$ | p |
| --- | --- | --- | --- | --- | --- | --- |
|  | N | % | N | % |  |  |
| **Parity** |  |  |  |  |  |  |
| Primiparous | 30 | 19.4 | 46 | 29.7 | 4.462 | **0.035** |
| Multiparous | 125 | 80.6 | 109 | 70.3 |  |  |
| **Time of childbirth** |  |  |  |  |  |  |
| At term | 123 | 79.4 | 132 | 85.2 | 6.548 | **0.038** |
| Preterm | 21 | 13.5 | 21 | 13.5 |  |  |
| Postterm | 11 | 7.1 | 2 | 1.3 |  |  |
| **Birth Type** |  |  |  |  |  |  |
| Vaginal birth | 55 | 35.5 | 47 | 30.3 | 0.935 | 0.334 |
| Cesarean section birth | 100 | 64.5 | 108 | 69.7 |  |  |
| **Labor process<sup>a</sup>** |  |  |  |  |  |  |
| Spontaneous | 26 | 17.2 | 24 | 15.5 | 1.716 | 0.424 |
| Oxytocin induction | 33 | 21.9 | 26 | 16.8 |  |  |
| Cesarean section | 92 | 60.9 | 105 | 67.7 |  |  |
| **Had any intervention in the second stage of birth** |  |  |  |  |  |  |
| Yes<sup>b</sup> | 22 | 14.2 | 21 | 13.5 | 0.027 | 0.869 |
| No | 133 | 85.8 | 134 | 86.5 |  |  |
| **Developed fetal distress in childbirth** |  |  |  |  |  |  |
| Yes | 13 | 20.6 | 0 | 0.0 | 11.659 | **0.001** |
| No | 50 | 79.4 | 50 | 100.0 |  |  |
| **Urgent puerperium hemorrhage status (observational)** |  |  |  |  |  |  |
| Mild hemorrhage | 70 | 45.2 | 100 | 64.2 | 12.217 | **0.002** |
| Moderate hemorrhage | 80 | 51.6 | 50 | 32.3 |  |  |
| Severe hemorrhage | 5 | 3.2 | 5 | 3.2 |  |  |

<sup>a</sup>Data of 4 participants could not be reached, <sup>b</sup>Episiotomy, Vacuum, Forceps, Fundal pressure, $\chi^2$ : Chi-squared test

In the logistic regression analysis, maternal obesity was found to increase the rates of multiparity by 1.758 times (1.038-2.978 %95 CI, p=0.036), post-maturity by 5.902 times (1.283-27.164 %95 CI, p=0.023) and moderate postpartum hemorrhage by 2.286 times (1.433-3.646 %95 CI, p=0.001) (Table 3).

**Table 3.**
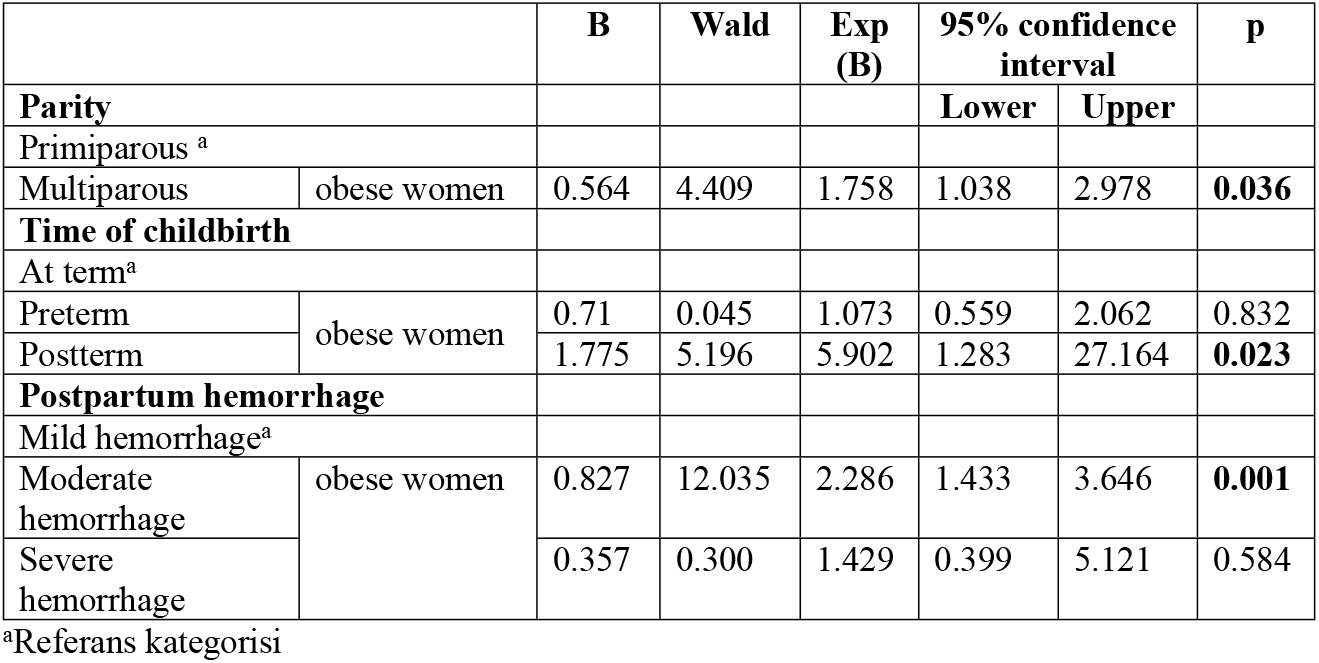
Regression analysis of some obstetric characteristics of women (N=310)

No interventional application was made to women during the data collection process. In addition, no direction was given that could lead to negative attitude and behavior changes. The study did not have any side effects or adverse effects on participants.

## Discussion

In this study, the effects of maternal obesity on some obstetric characteristics of women were examined. It was determined that the majority of the obese participants were multiparous and gave birth at term. Additionally, postterm delivery and moderate postpartum hemorrhage were observed more frequently in the obese group. Maternal obesity was determined to increase the rates of multiparity (1.758 times), post-maturity (5.902 times), and moderate postpartum hemorrhage (2.286 times). Fetal distress at birth developed only in the infants of the obese participants. There was no significant difference between the groups in terms of their modes of delivery, labor induction statuses, and statuses of undergoing intervention in the second stage of labor.

Studies have shown multiparity as one of the causes of weight gain in women of reproductive age. In most of such studies, a positive association between parity and weight gain or BMI has been reported [19-20]. In one study, it was determined that overweight women were more likely to be unable to lose the weight they gained during pregnancy than underweight women were [21]. Gestational weight gain was also found to negatively affect the return to pre-pregnancy weight and postpartum weight loss [19]. Parity was positively associated with long-term obesity risk among Chinese women [20]. A positive correlation was also observed between numbers of deliveries and obesity in Iranian women. In particular, it was shown that high parity was associated with BMI [22]. In this case, it can be stated that multiparous women have a higher risk of obesity.

Pregnancies lasting longer than 40 weeks were associated with maternal obesity and especially morbid obesity (*OR*:2.32, *95% CI*:1.73–3.12) [23]. Similarly, post-maturity was observed in obese women in a retrospective study in Russia [24]. Studies have reported higher rates of post-maturity in obese people [18, 25]. In one study, it was stated that the incidence of preterm childbirth was lower in obese women compared to non-obese women.^25^ In a systematic review, the risk of preterm birth was not found to be significantly different in infants whose mothers were overweight or obese [26]. In two cohort studies, however, the risk of preterm childbirth was found to be higher in overweight and obese women than in normal-weight women [27-28]. It was observed that the sample sizes of the aforementioned studies were quite large. These differences may be due to the differences among their sample sizes. Based on the information obtained, no clear conclusion could be reached about the relationship between preterm childbirth and obesity.

The likelihood of postpartum hemorrhage is moderately increased in obese women compared to women with normal BMI [29]. In a previous study, it was found that obesity did not increase the occurrence of severe hemorrhage [30]. In a study conducted in the Netherlands, obesity was not found to be associated with major obstetric hemorrhage[31]. In another study, it was reported that obesity did not affect postpartum hemorrhage status [32]. NICE (2017) emphasized that a BMI of 35 kg/m^2^ or above increases the risk of postpartum hemorrhage [33]. Meta-analysis studies have confirmed that a BMI of 25.00 kg/m^2^ or above increases the risk of postpartum hemorrhage [17, 34-35]. Similar to studies with high levels of evidence, postpartum hemorrhage was observed in the obese participants in this study, and it can be argued that obesity increases the risk of hemorrhage.

In a systematic review, infants whose mothers were overweight or obese were found to be at a higher risk of stillbirth than infants whose mothers were of normal weight [26]. Similarly, another study revealed that overweight or obese women have higher rates of stillbirth and neonatal death than those of normal weight [36]. Intrauterine hypoxia was observed in the infants of women with 3rd obesity [24]. The infants of obese women have a higher risk of developing fetal distress compared to the infants of normal-weight women (*OR*:2.67, *95% CI*:1.18–3.58) [37]. The results of other studies in the relevant literature and those of this study suggested the infants of obese women have a higher risk of developing fetal distress during birth.

In a previous study, there was no significant difference between obese women and normal-weight women in terms of their modes of delivery [30]. In a study conducted in China, it was stated that the rates of cesarean section deliveries were very high, and the increased prevalence of obesity in women was among the reasons that could explain the high cesarean rates [38]. In another study, increased BMI was associated with the likelihood of cesarean section. Overweight and obese women were found to have a 1.58 to 2.75-fold greater risk of cesarean section deliveries [39]. In this study, no significant difference was found between the normal weight and obese groups in terms of their modes of delivery. In our country, Cesarean section is perceived as a painless mode of delivery and is generally preferred by individuals with high socio-economic status. Since the institution where this study was conducted is a public hospital, usually individuals with low socio-economic status present to the institution. Moreover, cesarean delivery in the institution is performed based on necessity, not preference. There may not have been a difference between the groups due to the low socio-economic levels of the women included in the study and the fact that cesarean section was not optional.

One study showed that the rates of spontaneous labor initiation were higher than predicted in obese pregnant women. However, the authors determined that the rate of spontaneous labor was lower in obese women compared to normal-weight women. Moreover, in the same study, it was observed that the need for labor induction was more prevalent in obese women compared to normal-weight women [32]. In another study, BMI was found to not affect the rates of deliveries requiring intervention [39]. It was reported that as BMI increased, the rate of spontaneous labor decreased [40]. In a meta-analysis study, it was found that the incidence of vaginal childbirth requiring intervention was higher in obese women.

## Strengths and Limitations

There were some limitations in this study. The fact that the patient rooms in the maternity ward of the hospital where this study was conducted have 4 or more beds, and there are baby-sized beds next to the mothers’ beds may have caused the space to narrow and the room to be crowded and noisy. For this reason, there was difficulty in creating sufficient space and establishing communication while measurements were being made. Additionally, due to the COVID-19 pandemic that emerged during the data collection process, the access of health workers working in external units to the gynecology service was restricted, the families were more anxious, and the unit’s employees preferred their procedures to be performed with less contact. Women in the mildly obese category were not included in this study. Thus, the results might not apply to women who are mildly obese.

The strength of this study is that there a strong correlation was found between maternal obesity and some complications in childbirth and the postpartum period. The significance of these findings can be examined from different perspectives. The results indicate the need for advice and counseling programs for obese women to lose weight before pregnancy. They also point to the classification of obese pregnancies as high-risk pregnancies and the provision of appropriate antenatal care to these women. Additionally, they suggest that widespread obesity among women of reproductive age is associated with several health risks for women later in life.

## Conclusion

In this study, it was determined that the majority of the participants in the maternally obese group were multiparous and gave birth at term. However, postterm birth and moderate postpartum hemorrhage were found to be more common in the obese group. Fetal distress in childbirth developed only in the infants of the obese women. It is important to provide counseling or referral to a nutrition and dietetics specialist so that women who have obesity problems in the preconception period can reach normal BMI values. As the number of deliveries increases, the risk of obesity increases, and midwives should provide counseling to women about healthy nutrition and weight control during both the pregnancy and postpartum periods. Starting from the pre-pregnancy period until the end of the postpartum period, midwives have an important role in providing the necessary attention and care to mothers with obesity problems and regulating healthy lifestyle habits and behaviors. To prevent obesity, which is a health problem of our age, and the complications it causes and develop potential interventions, it is recommended to conduct extensive research on factors related to maternal obesity and make sense of the associated mechanisms.

## Data Availability

All relevant data are within the manuscript and its Supporting Information files.

## References

1. Centers for Disease Control and Prevention. Adult obesity facts [Internet]. Centers for Disease Control and Prevention: 2022. [cited 2022 July 30]. Available from: https://www.cdc.gov/obesity/data/adult.html#:~:text=Obesity%20is%20a%20common%2C%%20%2020serious,from%204.7%25%20to%209.2%25

2. Centers for Disease Control and Prevention. Disability and obesity [Internet]. Centers for Disease Control and Prevention: 2019. [cited 2022 July 29]. Available from: https://www.cdc.gov/ncbddd/disabilityandhealth/obesity.html#:∼:text=An%20adult%20who%20has%20a,or%20higher%20is%20considered%20obese

3. The American College of Obstetricians and Gynecologists (ACOG). Obesity and pregnancy [Internet]. The American College of Obstetricians and Gynecologists: 2021. [cited 2022 July 27]. Available from: https://www.acog.org/womens-health/faqs/obesity- and%20pregnancy#:~:text=Being%20overweight%20is%20defined%20as,BMI%20of%2030%20to%2034.9

4. Simko M, Totka A, Vondrova D, Samohyl M, Jurkovicova J, Trnka M, et al. Maternal Body Mass Index and Gestational Weight Gain and Their Association with Pregnancy Complications and Perinatal Conditions. Int. J. Environ. Res. Public Health 2019;16(10):1751. PMID: 31108864 https://doi.org/10.3390/ijerph16101751

5. National Institute for Health and Care Excellence (NICE). Obesity: identification, assessment and management [Internet]. NICE: 2014. [cited 2022 July 27]. Available from: https://www.nice.org.uk/guidance/cg189/ifp/chapter/obesity-and-being-%20%20overweight#:~:text=What%20BMI%20means%20for%20adults,40%20or%20more%20%E2%80%93%20severely%20obese

6. WHO. Obesity and overweight [Internet]. WHO: 2021. [cited 2022 July 31]. Available from: https://www.who.int/news-room/fact-sheets/detail/obesity-and-overweight

7. Hacettepe University Institute of Population Studies. 2018 Turkey Demographic and Health Survey [Internet]. Hacettepe University Institute of Population Studies, T.R. Presidency of Turkey Directorate of Strategy and Budget and the Scientific and Technological Research Council of Turkey (TÜBİTAK), Ankara, Türkiye: 2019. [cited 2022 July 27]. Available from: http://www.sck.gov.tr/wp-content/uploads/2020/08/TNSA2018_ana_Rapor.pdf

8. Hill B, McPhie S, Moran LJ, Harrison P, Huang TTK, Teede H, et al. Lifestyle intervention to prevent obesity during pregnancy: Implications and recommendations for research and implementation. Midwifery 2017;49:13–18. PMID: 27756642 https://doi.org/10.1016/j.midw.2016.09.017

9. Vitner D, Harris K, Maxwell C, Farine D. Obesity in pregnancy: a comparison of four national guidelines. J Matern Fetal Neonatal Med 2019;32(15):2580–2590. PMID: 29447091 https://doi.org/10.1111/j.14710528.2004.00546.x

10. The Royal College of Obstetricians and Gynaecologists (RCOG). Care of women with obesity in pregnancy (Green-top Guideline No. 72) [Internet]. The Royal College of Obstetricians and Gynaecologists: 2018. [cited 2022 July 27]. Available from: https://www.rcog.org.uk/guidance/browse-all-guidance/green-top-guidelines/care-of-women-with-obesity-in-pregnancy-green-top-guideline-no-72/

11. Luccisano SP, Weber HC, Murfet GO, Robertson IK, Prior SJ, Hills AP. An audit of pre-pregnancy maternal obesity and diabetes screening in rural regional tasmania its impact on pregnancy and neonatal outcomes. Int J Environ Res Public Health 2021;18(22):12006. PMID: 34831762 https://doi.org/10.3390/ijerph182212006.

12. Gaillard R. Maternal obesity during pregnancy and cardiovascular development and disease in the offspring. Eur J Epidemiol 2015;30(11):1141–1152. PMID: 26377700 https://doi.org/10.1007/s10654-015-0085-7

13. Marchi J, Berg M, Dencker A, Olander EK, Begley C. Risks associated with obesity in pregnancy, for the mother and baby: a systematic review of reviews. Obesity Reviews 2015;16(8):621–638. PMID: 26016557 https://doi.org/10.1111/obr.12288

14. Thompson AM, Thompson JA. An evaluation of whether a gestational weight gain of 5 to 9 kg for obese women optimizes maternal and neonatal health risks. BMC Pregnancy Childbirth 2019;19(1):126. PMID: 30975086 https://doi.org/10.1186/s12884-019-2273-z

15. Kiliç M, Erci B. The effect of the care provided based on self-care model of orem on self-care agency and frequency of nursing diagnoses in pregnant women with threat of preterm birth. Turkiye Klinikleri J Nurs Sci 2017;9(1):1–14. https://doi.org/0.5336/nurses.2015-49259

16. Batman D, Şeker S. The Effect of Web Based Education on the Level of Self-Confidence and Anxiety in Care of Parents of Premature Infants.DEUHFED 2019;12(2):107–115. https://dergipark.org.tr/en/pub/deuhfed/issue/54241/735027

17. Durmaz A, Komurcu N. Relationship between maternal characteristics and postpartum hemorrhage: a meta-analysis study. JNR 2018;26(5):362–372. PMID: 29219937 https://doi.org/10.1097/jnr.0000000000000245

18. Salameh K, Al-Bedaywi R, Habboub L, Elkabir NA, Tomerak A. Extremely Post-Term Infant with Adverse Outcome. Clin Res Pediatr 2018;1(2):1–4. https://doi.org/10.33309/2638-7654.010208

19. Rebholz SL, Jones T, Burke KT, Jaeschke A, Tso P, D’Alessio DA, et al. Multiparity leads to obesity and inflammation in mothers and obesity in male offspring. American Journal of Physiology-Endocrinology and Metabolism 2012;302(4):E449–E457. PMID: 22127227 https://doi.org/10.1152/ajpendo.00487.2011

20. Li W, Wang Y, Shen L, Song L, Li H, Liu B, et al. Association between parity and obesity patterns in a middle-aged and older Chinese population: a cross-sectional analysis in the Tongji-Dongfeng cohort study. Nutr Metab 2016;13(1):1–8. PMID: 27795732 https://doi.org/10.1186/s12986-016-0133-7

21. Rooney BL, Schauberger CW. Excess pregnancy weight gain and long-term obesity: one decade later. Obstet Gynecol 2002;100(2):245–252. PMID: 12151145 https://doi.org/10.1016/S0029-7844(02)02125-7

22. Hajiahmadi M, Shafi H, Delavar MA. Impact of parity on obesity: a cross-sectional study in Iranian women. Med Princ Pract 2015;24(1):70–74. PMID: 25402350 https://doi.org/10.1159/000368358

23. Woolner AM, Bhattacharya S. Obesity and stillbirth. Best Pract Re Clin Obstet Gynaecol 2015;29(3):415–426. PMID: 25457855 https://doi.org/10.1016/j.bpobgyn.2014.07.025.

24. Akhmetovna KL, Eduardovna TA, Vladimirovna CL, Vladislavovich MN. Metabolic disturbances in obese pregnant residents of an industrial region (The Urals, Russia). Oman Med J 2016;31(3):211. PMID: 27162592 https://doi.org/10.5001/omj.2016.40

25. Dixit A, Girling JC. Obesity and pregnancy. Journal of Obstetrics and Gynaecology 2008;28(1):14–23. PMID: 18259892 https://doi.org/10.1080/01443610701814203

26. Liu P, Xu L, Wang Y, Zhang Y, Du Y, Sun Y, et al. Association between perinatal outcomes and maternal pre-pregnancy body mass index. Obesity Reviews 2016;17(11):1091–1102. PMID: 27536879 https://doi.org/10.1111/obr.12455

27. Bacârea A, Bacârea VC, Tarcea M. The relation between prepregnancy maternal body mass index and total gestational weight gain with the characteristics of the newborns. J Matern Fetal Neonatal Med 2020;35(17):3284–3289. PMID: 32924693 https://doi.org/10.1080/14767058.2020.1818205

28. Sobczyk K, Holecki T, Woźniak-Holecka J, Grajek M.Does Maternal Obesity Affect Preterm Birth? Documentary Cohort Study of Preterm in Firstborns—Silesia (Poland). Children 2022;9(7):1007. PMID: 35883991 https://doi.org/10.3390/children9071007

29. Butwick AJ, Abreo A, Bateman BT, Lee HC, El-Sayed YY, Stephansson O, et al. Effect of maternal body mass index on postpartum hemorrhage. Anesthesiology 2018;128(4):774–783. https://doi.org/10.1097/ALN.0000000000002082

30. Pallasmaa N, Ekblad U, Gissler M, Alanen A. The impact of maternal obesity, age, preeclampsia and insulin dependent diabetes on severe maternal morbidity by mode of delivery—a register-based cohort study. Arch Gynecol Obstet 2015;291(2):311–318. PMID: 25115277 https://doi.org/10.1007/s00404-014-3352-z

31. Witteveen T, Zwart JJ, Gast KB, Bloemenkamp KW, van Roosmalen J. Overweight and severe acute maternal morbidity in a low-risk pregnant population in the Netherlands. PLoS One 2013;8(9):e74494. PMID: 24069316 https://doi.org/10.1371/journal.pone.0074494

32. Lauth C, Huet J, Dolley P, Thibon P, Dreyfus M. Maternal obesity in prolonged pregnancy: Labor, mode of delivery, maternal and fetal outcomes. J Gynecol Obstet Hum Reprod 2021;50(1):101909. PMID: 32927107 https://doi.org/10.1016/j.jogoh.2020.101909

33. National Institute for Health and Care Excellence. Intrapartum care: Care of healthy women and their babies during childbirth. NICE: 2017. Available from: https://www.nice.org.uk/guidance/cg190/evidence/full-guideline-pdf-248734770 Accessed: 2023 February 12

34. Higgins N, Patel SK, Toledo P. Postpartum hemorrhage revisited: new challenges and solutions. Curr Opin Anesthesiol 2019;32(3):278–284. PMID: 31045634 https://doi.org/10.1097/ACO.0000000000000717

35. Ende HB, Lozada MJ, Chestnut DH, Osmundson SS, Walden RL, Shotwell MS, et al. Risk factors for atonic postpartum hemorrhage: a systematic review and meta-analysis. Obstet Gynecol 2021;137(2):305. PMID: 33417319 https://doi.org/10.1097/AOG.0000000000004228

36. Yu YH, Bodnar LM, Himes KP, Brooks MM, Naimi AI. Association of overweight and obesity development between pregnancies with stillbirth and infant mortality in a cohort of multiparous women. Obstet Gynecol 2020;135(3):634. PMID: 32028483 https://doi.org/10.1097/AOG.0000000000003677

37. Frolova AI, Raghuraman N, Stout MJ, Tuuli MG, Macones GA, Cahill AG. Obesity, second stage duration, and labor outcomes in nulliparous women. Am J Perinatol 2021;38(04):342–349. PMID: 31563134 https://doi.org/10.1055/s-0039-1697586

38. Mi J, Liu F. Rate of caesarean section is alarming in China. Lancet 2014;383(9927):1463–1464. PMID: 24766963 https://doi.org/10.1016/S0140-6736(14)60716-9

39. Angeliki A, Dimitrios P, Chara T. Maternal obesity and its association with the mode of delivery and the neonatal outcome in induced labour: implications for midwifery practice. Eur J Midwifery 2018;2:4. PMID: 33537565 https://doi.org/10.18332/ejm/85792

40. Harper LM, Caughey AB, Odibo AO, Roehl KA, Zhao Q, Cahill AG. Normal progress of induced labor. Obstet Gynecol 2012;119(6):1113–1118. PMID: 22569121 https://doi.org/10.1097/AOG.0b013e318253d7aa

